# Sanitation barriers, lack of awareness and unaffordability affecting menstrual health management among urban low-income group, informal sector women workers: A study from Mumbai suburbs

**DOI:** 10.64898/2026.08.13.26360180

**Authors:** Sarah Shaikh, Sajal Sharma, Vruti Shah, Pooja Mehta, Madhuraka Pal

## Abstract

**Background and objectives:** Millions of women across the world experience menstruation, yet for those in the low-income, informal urban workforces, it can be accompanied by an unpleasant experience due to poor sanitation, financial constraints and lack of awareness. In the Indian urban context, despite a decade of implementing policy initiatives, including the Rashtriya Kishor Swasthya Karyakram and the Menstrual Hygiene Scheme by the Government of India, urban women of reproductive age in the low-income group informal workforce have seen limited benefits. We sought to understand the gaps preventing healthy menstrual health management (MHM) among these women in Mumbai’s suburbs and aimed to learn, directly from them, what interventions might help in their MHM.

**Methods:** 100 randomly selected women of reproductive age, belonging to the low-income group urban work force, in Mumbai Suburbs were surveyed. A structured 18-item questionnaire was used to conduct a one-on-one interview, surveying about demographics, menstrual hygiene product use, hygiene practices, sanitation access, diet, and challenges faced.

**Results:** More than half the participants of the study (53%) earned under INR10,000 a month and 87% were aged above 18 years. A striking 79% of the participants had experienced at least one clinically significant menstrual morbidity; sanitation barriers affected 67% while 15% were forced to change pads in unsafe, open spaces; and adequate nutrition during periods was reported by only 24%. 57% of the participants reported that a combination of awareness, better toilets and subsidised pads will help them manage their menstrual hygiene better.

**Interpretation and conclusions:** For low-income women in the informal urban workforce, menstrual health challenges arise from gaps in knowledge, inadequate sanitation infrastructure and unaffordability of menstrual hygiene products. This highlights the need for coordinated action-spreading awareness, building accessible sanitation facilities, and strengthening primary healthcare to screen and treat menstrual morbidities- along with distribution of subsidised menstrual hygiene products for these underserved population.

## Introduction

Nearly half of the world’s population experience menstruation at some point in their lives. Menstruation, often referred to as "period," is the regular, monthly discharge of blood and tissue from the uterus through the vagina as a part of the reproductive cycle of girls and women of reproductive age. Menstrual health is defined as a state of complete physical, mental, and social well-being and not merely the absence of disease or infirmity in relation to the menstrual cycle^1,2^. Millions of menstruating girls and women worldwide experience period poverty, described as limited access to menstrual education, period products, or adequate water sanitation and hygiene (WASH) facilities^3^. Nationally representative government survey data show that although 90 per cent of urban Indian women aged 15–49 yrs use hygienic methods of menstrual protection compared to 73 per cent of their rural counterparts, more than half of the women in this age group remain anaemic nationally, indicating that urban residence alone does not guarantee menstrual health equity^4^. Menstrual morbidities and associated physical symptoms ranging from, weakness and pain to severe anaemia and Reproductive Tract Infection (RTI) make the experience of period difficult for a large fraction of women of reproductive age and the use of hygienic menstrual materials have shown to significantly lower the odds of self-reported RTI ^5^. Lack of access to adequate water sanitation and hygiene, proper nutrition, quality menstrual hygiene products and lack of awareness further compound these difficulties for women, especially for the ones who belong to the low-income group of the socio-economic structure of the society. In India, despite significant government and national policy initiatives such as the Rashtriya Kishor Swasthya Karyakram (RKSK) and the Menstrual Hygiene Scheme (MHS), the translation of policy into improvement in lives of females has been uneven, especially for adult women in urban informal settlements^6,7^, who comprise the India’s vast informal economy sector—as domestic helpers, janitors, and daily-wage workers.

The University Grants Commission (UGC) introduced ‘Fostering Social Responsibility and Community Engagement in Higher Educational Institutions’ as an integral component of higher education, in accordance with the recommendations of the National Education Policy (NEP) 2020, which envisages Higher Educational Institutions (HEIs) as significant contributors to India’s socio-economic development through active community engagement^8^. The objective is to bridge the gap between classroom learning and social realities by encouraging closer relationship between HEIs and local communities to identify and address issues encountered by communities in their daily lives. In accordance with the UGC guidelines, SVKM’s Mithibai College of Arts, Chauhan Institute of Science and Amrutben Jivanlal College of Commerce and Economics, Mumbai, has collaborated with the Unnat Maharashtra Abhiyan, coordinated by IIT Bombay, to institutionalize the Community Engagement Program (CEP). From an academic perspective, the CEP is structured as a mandatory two-credit course offered to the second-year undergraduate students across all disciplines, with strong emphasis on experiential and field-based learning. The course comprises a total 30 hours of teaching-learning, with 8 contact hours devoted to orientation, guided support for topic selection, study design, survey preparation under faculty mentorship. The remaining 22 hours are assigned to field work, data collection, analysis and report writing, ensuring active student participation and meaningful interaction with local communities, industries, artisans and small businesses.

As a part of this initiative, students of Mithibai College, Mumbai, conducted this study by reaching out with a structured questionnaire for survey about menstrual hygiene, practices and awareness to women from lower-income groups representing a significant section of the urban informal workforce in suburban areas of Mumbai. Although educational qualification was not included in the questionnaire, informal inquiry about the respondents’ education qualification revealed that the respondents ranged from illiterate to those who had completed primary, secondary, and even higher secondary education. Several respondents were unable to read or understand the questionnaire due to limited literacy and each question was explained to them in Hindi/ Marathi, aiding them in recording their responses. Based on these interactions, they were advised on adopting safer practices, such as using sanitary pads instead of cloth, to consult a doctor in cases of severe or frequent menstrual pain, etc. This study was aimed at understanding the lacuna that acts as a hurdle for the women of the low-income group informal work force in the Mumbai suburban region in experiencing a healthy period and in proper MHM, along with finding out what interventions would help these women manage their menstrual health better.

## Materials and methods

### Area of Study

The study was conducted in the Mumbai Suburban Region namely Juhu/Vile Parle, Andheri and Borivali and was focused on women belonging to lower-income groups, including housewives, domestic helpers, janitors, and cleaning staff, who comprised the informal workforce in Mumbai. These groups were selected because they are more likely to face financial constraints, frequently residing in overcrowded settlements characterised by shared sanitation, intermittent water supply and limited privacy, and insufficient guidance regarding menstrual hygiene management. Such socio-economic and infrastructural challenges directly influence their awareness, practices, and overall menstrual health experience.

### Target Population

The target population consisted of women of reproductive age from low-income households, comprising the working age group according to the 2011 census, which is 15-64 years and were engaged in informal occupations. In our study, these women were grouped under four age categories: below 18 years, 18–26 years, 26–35 years, and above 35 years. This categorization helped in understanding how age, education, income, work environment, and family background affect menstrual hygiene management from adolescence to adulthood.

### Ethical approval and consent of participants

The ethical approval for the study was “waived” by the institutional ethical review committee. The study was a part of the Undergraduate program’s Community Engagement and Service curriculum, wherein students engage with local community which help them understand the local development needs and problems of immediate communities. Students are required to generate a report of their interaction and the findings for evaluation. In the current study, the students approached women of reproductive age in the aforementioned localities and verbally explained to them the content and the research and academic intent of the survey. The women who consented verbally to the content and the research and academic intent of the survey participated in the study and their responses were recorded by one of the students, while another student was a witness to the process. For the participants who were <18 years of age, verbal consent of participation from their mother/ female guardian was obtained upon explaining to them the content and the research and academic intent of the survey. Identities of the respondents were kept confidential and respondents were allowed to report their responses anonymously.

### Research Instrument

A structured physical questionnaire included 18 simple and clear questions related to demographic details, menstrual hygiene habits, sanitation and water access, dietary habits– general and during menstruation, health concerns, and challenges faced by the respondents. The questionnaire was piloted with 10 women outside the study area and minor revisions were made to improve clarity. Since many respondents belonged to less-educated backgrounds, the questions were explained in simple language to ensure proper understanding. Responses were collected through face-to-face interaction, which helped build trust and encouraged participants to share their real-life experiences.

### Sample Size

Given the time constraints of the CEP under the University of Mumbai curriculum, a total sample size of 100 respondents was selected randomly from the lower-income groups in the aforementioned localities in Mumbai.

### Data Collection Method

Primary data were collected through in-person interviews using the structured questionnaire comprising 18 questions between the dates 20/01/2026 and 30/01/2026 (both dates inclusive). Face-to-face interaction allowed respondents to express themselves comfortably in their preferred language and ensured clarity in responses. Secondary data were obtained from research papers, academic journals, and published reports related to MHM. These sources provided background information and supported the interpretation of primary findings.

The questions asked to the participants of the study were broadly divided under six categories (with multiple responses being allowed for some questions) to get information on the following:

1. Demographic profile of the participants.

2. Menstrual hygiene product used by the participants, the factors related to the selection of the product used and the difficulties faced in procuring the products.

3. Menstrual hygiene practices of the participants.

4. Food habits of participants and period fatigue experienced by participants.

5. Difficulties and challenges faced by respondents during their period.

6. According to the participants, what can help them manage their period better?

After the completion of the survey, the responses were plotted in MS excel to generate bar graphs and Venn diagrams, to comprehend the findings of the survey.

## Results and Discussion

### Demographic profile of the participants

The current study is targeted to the women of reproductive age in the low-income group (i.e. monthly family income < 50,000.00), constituting the informal workforce in the Mumbai Suburban area. Three questions were asked regarding demographics, particularly, their age, the monthly income and the occupation of the participants. Regarding the age of the participants, the modal group was 26–35 years (35%), followed by above 35 years (28%), 18–25 years (24%), and below 18 years (13%). Taken together, 63% of respondents were aged 26 years or above, representing women at peak reproductive and occupational burden (Figure 1A). Occupationally, 39% of respondents were maids or domestic helpers, 27% were housewives, 19% were janitors/cleaning staff, and 15% were other daily-wage workers (Figure 1B). Thus, 73% of the participants of the study were engaged in active physical labour outside the home with roles that typically involve commuting, prolonged work hours and minimal control over sanitation access. The monthly income was low across the cohort: 24% earned less than 5,000 and 29% earned 5,000– 10,000, placing 53% of the participants in a monthly income bracket below the income of 10,000. A further 23% earned 10,000– 15,000, and 24% earned above 15,000 monthly (Figure 1C), which made the participants socio-economically members of the low-income group (LIG), with total family income of less than 50,000.00 a month.

**Figure 1.**
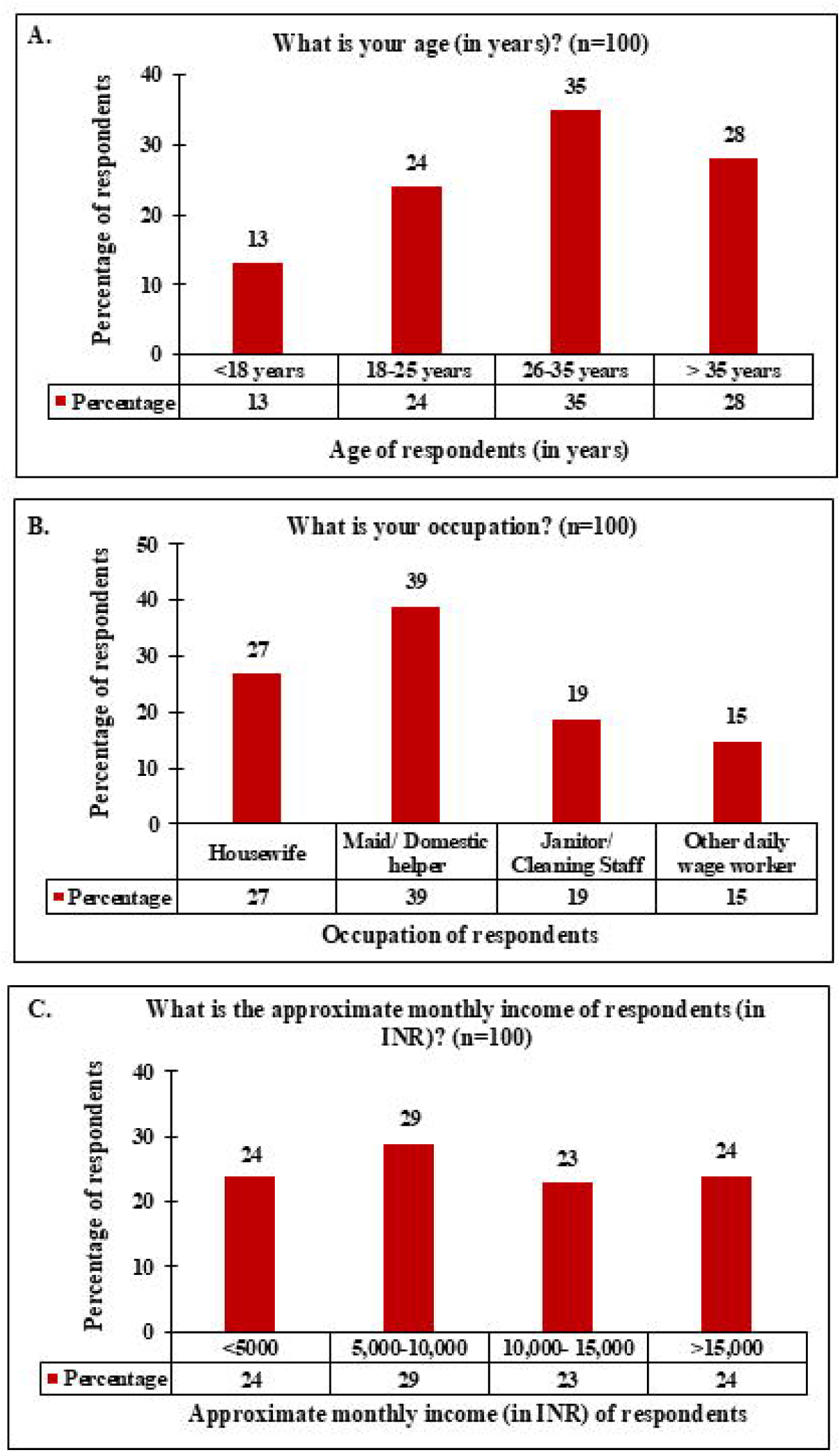
Demographic profile of survey respondents: (A) Bar graph representing the “Age distribution of respondents” across four categories (n=100); (B) Bar graph representing the “Occupation of respondents” (n=100); (C) Bar graph representing the “Approximate monthly income (in INR) of respondents” (n=100).

### 2. Menstrual hygiene product used by the participants and the factors influencing the selection of the menstrual hygiene product used

For this, four questions were asked to the cohort to get information on the choice of menstrual hygiene products used, the amount of money they spent every month on menstrual hygiene products, the factors that were responsible for the choice they made and any difficulties if they faced in procuring menstrual hygiene products. The most commonly used menstrual hygiene product by the cohort was local or cheap disposable pads (42%), followed by branded pads (29%), cloth (23%), and reusable pads (6%) (Figure 2A). 71% of the participants had transitioned to some form of disposable pads but 29% of the cohort used reusable pads or cloth as absorbent, indicating access to quality disposable menstrual hygiene products remained a concern for a significant number of the respondents. Cloth and reusable pad usage have disadvantages as they can cause severe health concerns, if not washed/ cleaned properly^9^. The monthly expenditure on menstrual products by the cohort ranged widely: 18% spent less than 50, 24% spent 50– 100, 35% spent 100– 200, and 23% spent above 200 (Figure 2B). When contextualised against the income distribution (53% earning below 10,000 per month), even the modal expenditure range of 100– 200 per month represents a disproportionate financial burden on the cohort, with the monthly expenditure on menstrual hygiene products being 2% or more of the monthly income in some cases. The cohort were asked about who decided the menstrual hygiene products they used and it was found that the decision-making autonomy over menstrual hygiene products was fragmented. 41% of the participants made independent decisions, 25% were influenced by family members, 12% by their husband, and 25% stated that their choice depended on the available funds (Figure 2C). Therefore, a staggering 58% of the women lacked full autonomy in decision-making regarding their own menstrual health, with money being the limiting factor in decision making for 25% of the women. The barriers to procuring menstrual hygiene products included lack of money (31%), shame or fear of social judgment (21%), and non-availability or distance from shop (23%). A total of 36% of respondents reported “no difficulty”—underscoring that a substantial majority (64%) encountered at least one access barrier with “money” being the limiting factor to access to menstrual hygiene products to 31% of the respondents, followed by 23% of the respondents having difficulty of access to menstrual hygiene products due to “distance/ non availability” (Figure 2D and 2E). Thus, “money” was a key factor in not just the decision making in “choosing” a menstrual hygiene product, but also on the “procurement” of menstrual hygiene products for about a fourth to a third of the respondents. This mirrors national spatial-analysis evidence identifying household wealth, education, caste, place of residence and toilet access as significant determinants of exclusive use of hygienic versus unhygienic menstrual products among young Indian women.^10^

**Figure 2.**
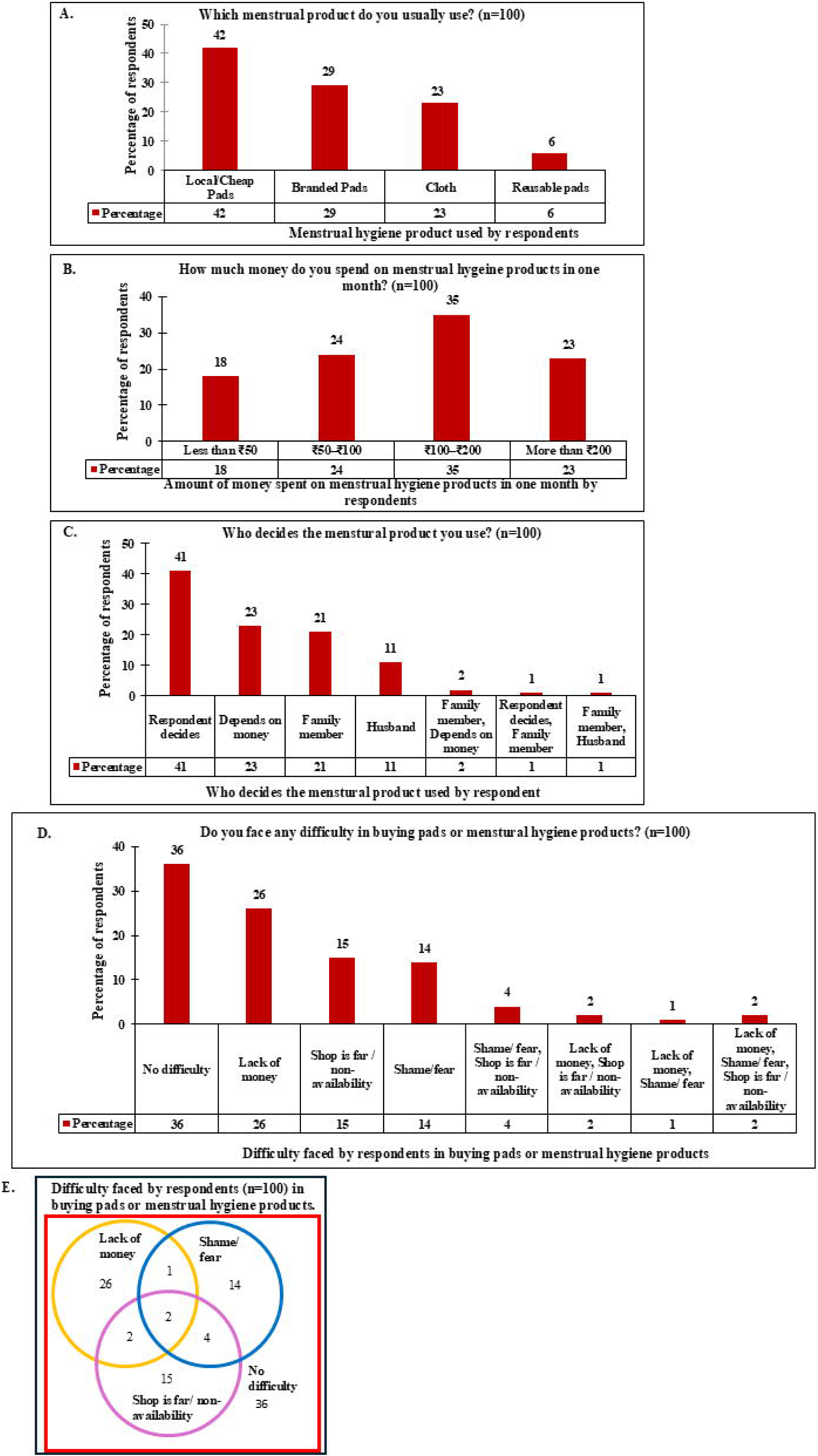
Menstrual hygiene products used, factors influencing the decision and difficulties faced by respondents in procuring the menstrual hygiene products: (A) Bar graph representing the choice of menstrual hygiene product most commonly used by respondents (n=100); (B) Bar graph representing the monthly expenditure on menstrual hygiene products by respondents (n=100); (C) Bar graph representing the “Decision-making factors for the menstrual hygiene product of choice used by respondents” (n=100); (D) Bar graph representing the “Difficulties faced in procuring menstrual hygiene products used by respondents” (n=100; multiple responses permitted); (E) Venn diagram of overlap of the “Difficulties faced in procuring menstrual hygiene products used by respondents” (n=100).

### 3. Menstrual hygiene practices of the participants

Next, three questions were asked to the respondents related to menstrual hygiene practices: “How often do you change your pad/ cloth in a day?”, “Where do you usually change your pad/ cloth?” and “Disposal method of used menstrual hygiene product”. Regarding pad/ absorbent change frequency (Figure 3A), 14% changed once daily and 35% changed twice daily— meaning 49% of the respondents, or nearly half, changed their products at or below twice per day, considerably lower than the recommended three-to-four changes in a day^11^. Only 24% met the recommended standard of changing absorbent/ pad three times or more, per day. Lesser frequency of changing absorbent can put women at potential risk of reproductive health problems, increasing the incidence of reproductive health morbidities and/ or RTI. The means of disposal of the menstrual waste products is also important. Kaur et al., 2018, discussed the importance of healthy menstrual hygiene practices and suggested of making girls aware of how to properly dispose of used menstrual products at home and in schools^12^. They also suggested of making girls aware of the consequences of throwing the menstrual waste in open or flushing them in toilets, reiterating the significance of proper menstrual waste disposal. Since, menstrual waste disposal is an important aspect of menstrual health management (MHM), we asked the respondents how they disposed their menstrual waste. Pad or cloth disposal practices by the respondents included wrapping and disposing in dustbins (41%), giving for municipal waste collection (19%), washing and reusing (25%), and disposing in open areas or drains (15%) (Figure 3B). Thus, less than half of the respondents (41%) were disposing their menstrual waste in a proper manner, upon wrapping. As per Kaur et al., 2018, reusable cloth/cloth pads should be hygienically cleaned and dried in sunlight for sterilization, followed by storage in clean and dry places avoiding contamination^12^. If this procedure is not stringently followed, a significant number of women of the 25% respondents who “wash and reuse” their menstrual hygiene products can be at a potential risk of RTI. The 15% of respondents using “open disposal” represent both a personal health risk and an environmental concern. The participants’ response to “Where do you usually change your pad/ cloth?” demonstrated that access to a private toilet was limited to the respondents and only 33% changed their absorbent in a private home toilet. A substantial 28% of the respondents used shared community toilets, 24% used workplace or school toilets, and 15%—the most vulnerable cohort—were compelled to use open or unsafe spaces (Figure 3C). These findings suggest that more awareness regarding menstrual hygiene and menstrual waste disposal need to be spread among women comprising the informal urban and metropolitan workforce along with building infrastructure for gender segregated toilets to provide privacy to women and ensure safety.

**Figure 3.**
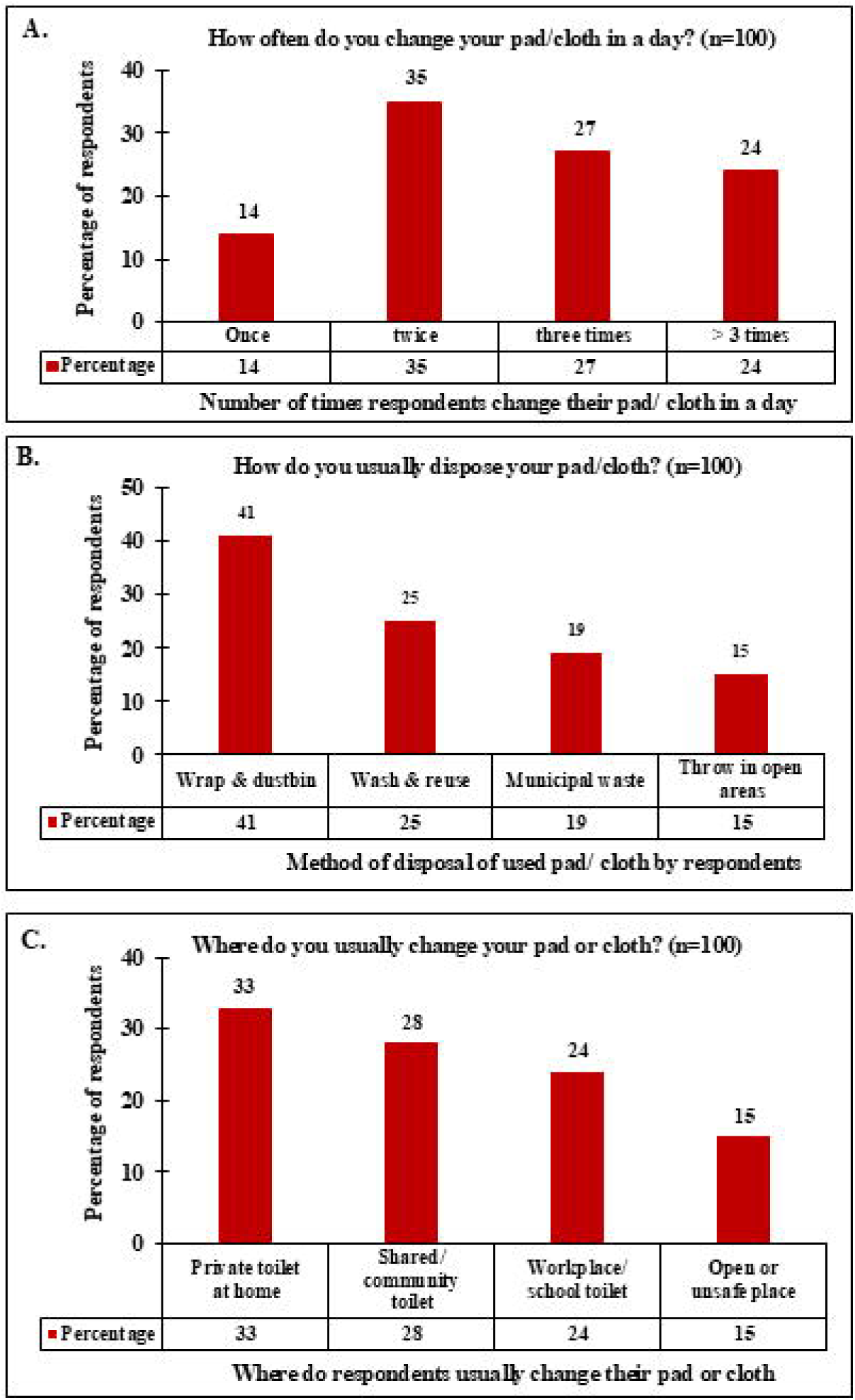
Menstrual hygiene practices of respondents: (A) Bar graph representing the “Frequency of changing pad/ cloth (absorbent) per day by the respondents” (n=100); (B) Bar graph representing the “Method of disposal of used menstrual hygiene products by the respondents” (n=100); (C) Bar graph representing the “Usual location for changing pad/cloth (absorbent) by the respondents” (n=100).

### 4. Food habits of respondents and period fatigue experienced by participants

Multiple studies have established the relationship between food habit and the occurrence and severity of period fatigue. Naraoka et al., 2023,^13^ investigated and discussed the role of nutrition and life-style habits in menstrual pain and period fatigue. They found that inadequate animal protein and vitamins in diet were related to sever dysmenorrhea in the study population of Japanese women of reproductive age. Munro et al., 2023,^14^ discuss the association between iron deficiency, anaemia and menstrual bleeding in women of reproductive age. Moschonis et al., 2013,^15^ observed that iron depletion in young girls was associated with high calcium intake, high consumption of fast foods, and low consumption of poultry and fruits. Thus, food intake and dietary habits played a very important role in menstrual morbidities, especially with anaemia, weakness and dizziness, all of which comprise period fatigue^16^. For this reason, we asked the respondents about their food habits and the physical challenges/ period fatigue faced by them during their period. Respondents were asked if they experienced tiredness, weakness, or dizziness during their period, and what was the frequency of the experience. It was found that the physical symptoms during menstruation were highly prevalent among the respondents: 36% experienced tiredness, weakness, or dizziness every month; 37% experienced them sometimes; 19% experienced them rarely; and only 8% never experienced them (Figure 4A). In aggregate, 73% reported regular or intermittent menstrual morbidity. It was also found that nutritional adequacy during menstruation was poor. During period, only 24% of the respondents reported proper and sufficient food intake; 42% relied predominantly on simple food (rice or roti and dal); 22% ate less than usual; and 12% frequently skipped meals entirely (Figure 4B). Combined, a staggering 76% of respondents experienced some degree of dietary inadequacy during their period. To inquire about iron and protein consumption among the respondents, we asked them how often they consumed green leafy vegetables, jaggery, dates, nuts, eggs, etc. Iron-rich food and proteins consumption— which are critical determinant of menstrual health given blood-loss-related iron depletion—was insufficient across the sample. Only 17% of the respondents consumed them daily; 36% consumed them three to four times per week; 32% consumed them once weekly; and 15% rarely or never consumed them (Figure 4C). Thus, 47% of the respondents, or nearly half of the sample, consumed iron-rich and protein rich foods once per week or less, making it a question of serious health concern.

**Figure 4.**
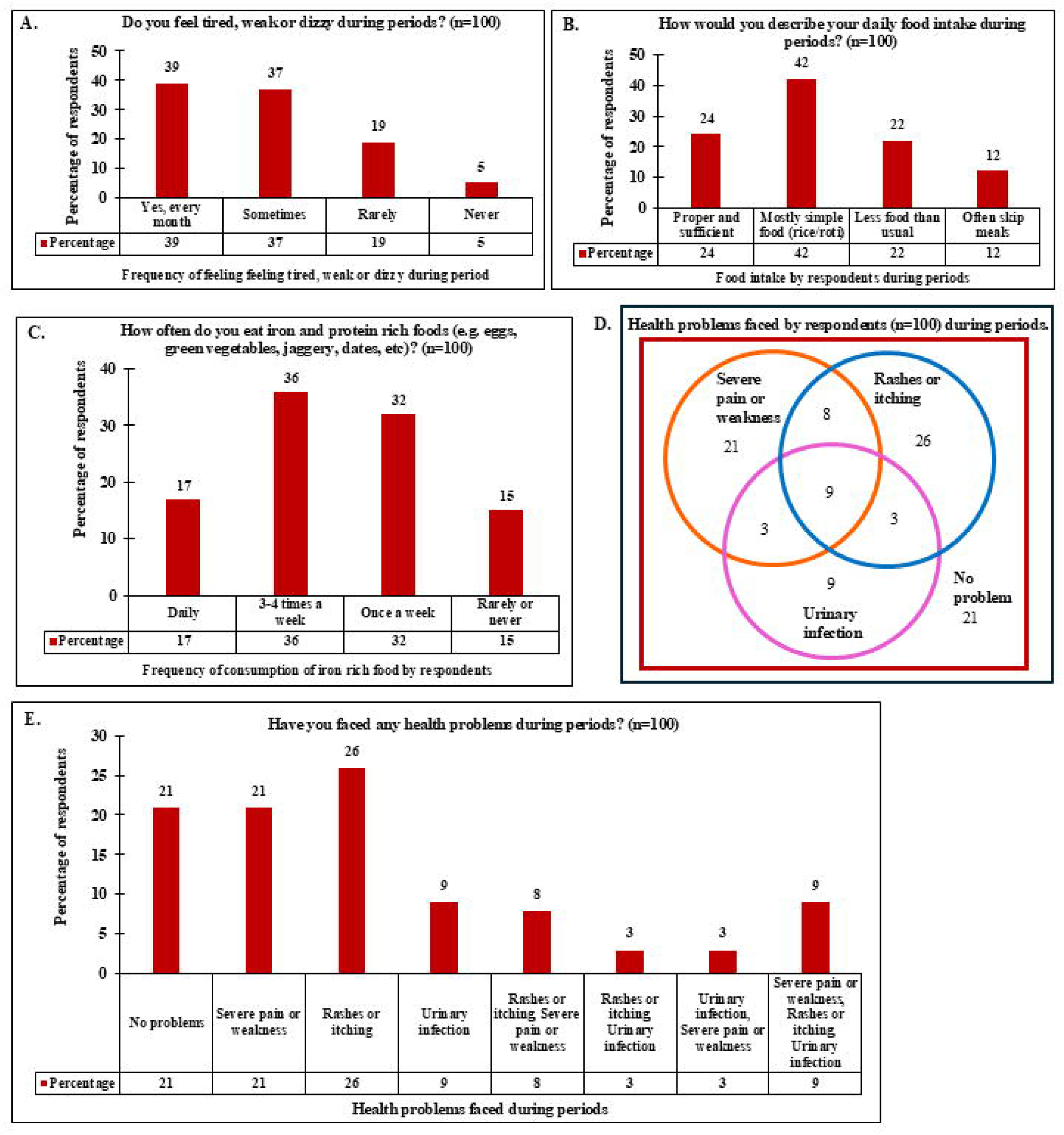
Food habit of respondents, menstrual fatigue and menstrual morbidities experienced by respondents: (A) Bar graph representing the “Frequency of feeling tired, weak or dizzy during periods by respondents” (n=100); (B) Bar graph representing the “Daily food intake during periods by respondents” (n=100); (C) Bar graph representing the “Frequency of consuming iron- and protein-rich foods (green leafy vegetables, jaggery, dates, nuts, eggs) by the respondents (n=100); (D) Venn diagram of overlap of “Health problems experienced by respondents during periods” (n=100); (E) Bar graph representing the “Health problems experienced by respondents during periods” (n=100; multiple responses permitted).

We also inquired about the health problems faced by the respondents during their period. Health problems during menstruation that were inquired about in the survey included severe pain or weakness, rashes or itching and urinary tract infections (multiple responses were permitted) (Figure 4D and 4E). The respondents revealed that 79% of the respondents experienced at least one clinically significant menstrual health issue. The respondents reported that 46% experienced rashes or itching, 41% suffered severe pain or weakness, 24% got urinary tract infections, and only 21% reported that they faced “no problems” during their period. The 79% of respondents, who experienced at least one clinically significant menstrual health issue, also mentioned informally that they relied on some pain killers or homemade remedies to alleviate the health challenge faced instead of seeking professional help to undergo proper medical examination and treatment. They also mentioned that they had never gotten their hemoglobin levels checked. 23% of the respondents experienced more than one of the three health problems in question during their period, and this fraction of the respondents might need immediate medical attention. Thus, there is an urgent need of spreading awareness about menstrual health among the low-income group female informal workforce in the urban and metropolitan regions along with the facilitation of provision of access to professional medical help, anemia screening and treatment of menstrual morbidities, at minimal cost.

### 5. Difficulties and challenges faced by respondents during period regarding MHM

Next, we asked the respondents about the problems they faced regarding their MHM and any societal/ other constraints they faced during their periods. For this we asked three questions and multiple responses were allowed for these questions: “What problems do you face the most related to toilet and clean water access during your period?”; “What restrictions (religious/ societal/ movement/ sleep) do you face during your period?” and “What problems do you face the most during periods?”

In this regard, we asked them about access to clean toilet, clean water and to privacy for managing the menstrual health during their periods. Only 33%, or a third of the respondents reported that they faced “no problem” in managing their menstrual health with respect to access to clean toilets, clean water and privacy. 38% respondents reported lack of clean toilets and 38% reported lack of clean water—the two most prevalent problems for the population under consideration—while 21% cited lack of privacy (Figure 5A). This 21% fraction of respondents reporting lack of privacy as a hindrance in their MHM encompass the 15% females who were forced to change their pads/ absorbent during their period in “open or unsafe” places along with some of the women who did not have access to gender segregated toilets at work/ dwelling areas. Taken together, a staggering 67% of the respondents faced at least one critical sanitation barrier during their period. In our opinion, allocation of funds for public health policies to build more gender segregated toilets with accessibility of clean water round the clock in or near dwelling areas, work places, bus stops, railway stations, etc. will help a larger number of women in the informal work force of metropolitan and suburban region of Mumbai to better manage their menstrual health. Such policies should also be extended to other cities and states as the lower income group informal workforce women in other cities and states might be facing these very problems in managing their menstrual health. The “Swaccha Bharat Abhiyan” has been a game changer in this aspect by giving people access to private toilets in their homes, but still more work is required on this front^17^. To make MHM of women hurdle free across the length and breadth of the nation and societies, it is required to allocate adequate funds for sanitation infrastructure with keeping the target population of low-income group women informal work force of the nation in mind.

**Figure 5.**
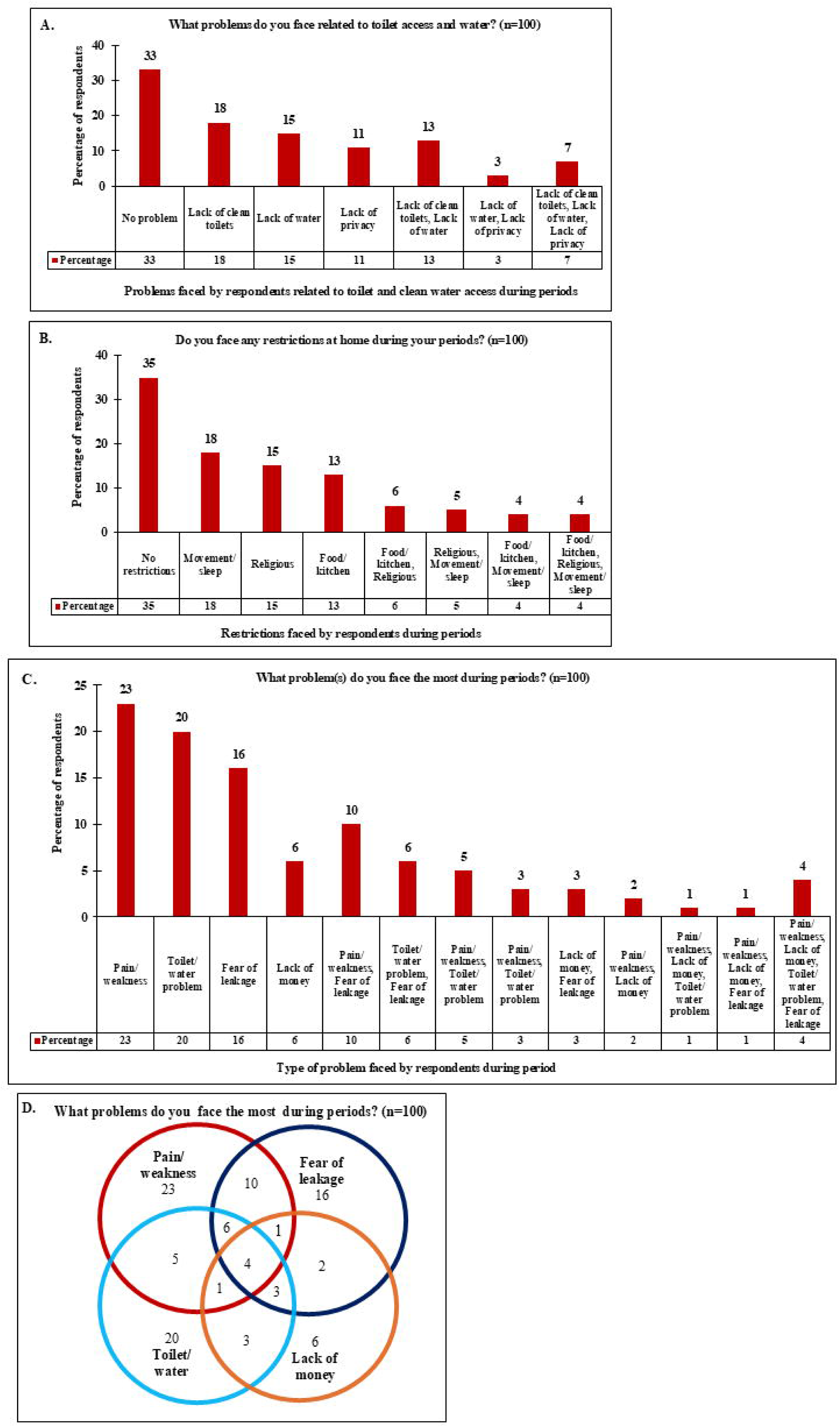
Sanitation barriers, restrictions, problems and challenges faced by respondents during periods: (A) Bar graph representing the “Problems related to toilet and clean water access (sanitation barriers) faced by respondents during period” (n=100; multiple responses permitted); (B) Bar graph representing the “Restrictions experienced at home during menstruation by respondents” (n=100; multiple responses permitted); (C) Bar graph representing the “Challenges (problems) perceived by respondents as most significant during periods” (n=100; multiple responses permitted); (D) Venn diagram illustrating the overlap among the “Problems/ challenges faced by respondents as the most significant during periods” (n=100).

**Figure 6.**
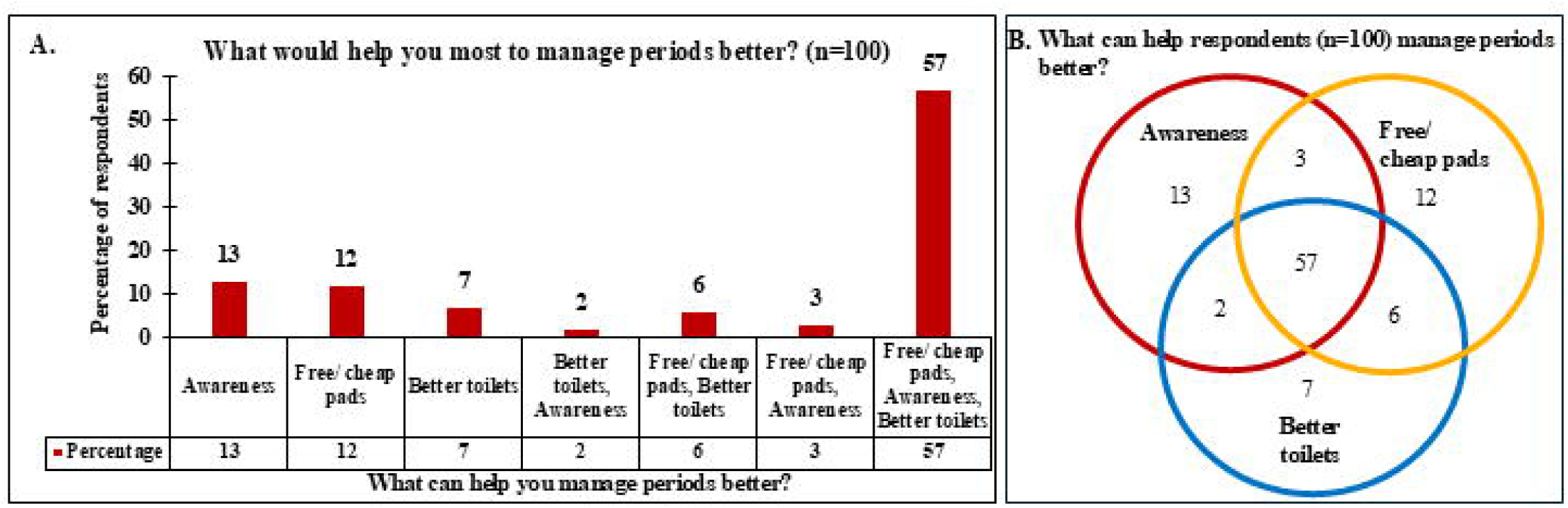
Interventions identified by respondents as most helpful for managing their menstrual health: (A) Bar graph of responses representing “What would help the respondents manage their period better?” (n=100; multiple responses permitted). (B) Venn diagram of the overlap between the three interventions that would help the respondents manage their period better (n=100).

To the question “What restrictions (religious/ societal/ movement/ sleep) do you face during your period?”, where multiple responses were allowed, the participants responded that “home restrictions” during menstruation were experienced by 65% of the respondents. “Movement and sleeping restrictions” affected 31%; religious restrictions were experienced by 30%; “food or kitchen restrictions” were reported by 27%; and 35% experienced no restrictions (Figure 5B). These responses show that low-income group women face societal as well as physical restrictions which could impact their MHM. Next, we asked the respondents about “the challenges they faced the most during their period”, encompassing pain, problems in toilet/water access, fear of staining and lack of money for menstrual hygiene product. Multiple responses were allowed and the most common problem faced during menstruation by the respondents were pain or weakness (49%) followed by fear of staining or leakage (45%), toilet or water problems (39%) and lack of money for products (17%) (Figure 5C and 5D). Thus, nearly half of the respondents experienced pain or weakness that impacted their lives in a negative way during their periods. Fear of staining or leakage experienced by 45% of the respondents could stem from lack of access to quality menstrual hygiene products and from the inability to change absorbent more frequently during periods as 49% of the respondents changed their products twice or below per day instead of changing absorbent at least three times a day. As opposed to the responses to the question “What problems do you face the most related to toilet and clean water access during your period?”, where 67% of the respondents reported that they faced at least one critical sanitation barrier, 39% of the respondents identified “Toilet/ water problems” as a major challenge in their MHM, reiterating the importance of sanitation infrastructure creation for access to the low-income group women.

### 6. Interventions that can help respondents manage their menstrual health better

Next, we asked the respondents that amongst “awareness”, “better toilets” and “free/ cheap (subsidized) pads”, “What can help them manage their period better?” Selection of multiple options were allowed and 57% of the respondents reported that all three, i.e. awareness, better toilets and free/ cheap pads, can help them manage their menstrual health better. This response corroborates with the other responses recorded in the survey. In the survey, 79% of respondents reported that they experienced at least one clinically significant menstrual health issue and mentioned informally that they did not seek professional help to undergo proper medical examination and treatment for the problems they faced; 15% of respondents used “open disposal” of their used menstrual hygiene products; only 24% of the respondents reported proper and sufficient food intake during their period and only 8% of the respondents never experienced tiredness, weakness, or dizziness during periods. These demonstrate that there was clearly a lack of knowledge and awareness, especially in terms of nutrition, menstrual morbidities, proper disposal of menstrual waste and other aspects of menstrual hygiene, which explains why 75% of the respondents report that “awareness” will help them manage their periods better. This lack of knowledge and awareness of how to manage their menstrual health and how to mitigate menstrual morbidities among the low-income group women constituting the informal work force in Mumbai suburbs, most likely, extrapolate to the low-income group women in other parts of the nation and this problem would require immediate attention.

In relation to the questions asked about the sanitation challenges the respondents faced, 67% of the respondents reported that they faced at least one critical sanitation barrier, 15% respondents were compelled to use open or unsafe spaces to change pads/ absorbents and 39% of the respondents identified “Toilet/ water problems” as a major challenge in their MHM, correlating to the number of respondents (72%) believing that “better toilets” will help them manage their periods better. Regarding the financial burden and difficulties in procuring quality menstrual hygiene product in question, 53% of the participants in the study had a monthly income below 10,000; 58% respondents had a monthly expenditure of > 100.00 a month for menstrual hygiene products, ; 22% respondents reported that the “shop was far away” and 31% respondents reported that “lack of money” were major barriers in procuring pads/ menstrual hygiene products; 25% of the respondents reported that the menstrual hygiene product they used “depends on the money” corroborates with 78% participants responding that “free/ cheap pads” would help them manage their menstrual health better.

Thus, the women who participated in this study, clearly articulated the need for an integrated intervention that include product affordability, sanitation infrastructure, and spreading of awareness regarding menstruation and menstrual health to transform the natural process of menstruation into a healthy “period” for them. The Government of Maharashtra’s Asmita Yojana^18^ and Supreme Court of India’s January 30, 2026 verdict^19^ recognized menstrual health as integral component of individual fundamental right under Article 21 (Right to Life and Dignity) highlights the growing attention to this issue. However, both these initiatives target the school going girls and rural women, leaving the women of reproductive age in the low-income group informal workforce in urban areas out of its’ coverage^20^.

For policymakers, this should translate into a mandate for holistic, equity-centered MHM and framing of menstrual hygiene policy that recognize menstrual health not as a personal hygiene matter, but as a fundamental issue of gender equity, public health and social development. The responses given by the participants of the survey not only point at building of sanitation infrastructure accessible to the women of low-income group informal workforce, but also to the building of more Primary Health Centers (PHCs) for treatment of menstrual morbidities and anaemia in these women. The PHCs can also be instrumental in distribution of menstrual hygiene products to these women at subsidised rates along with helping spread awareness about menstrual hygiene amongst these women. There is also a need for policies that facilitate checking hemoglobin levels of women at least biannually at minimal cost at the PHCs. The policy makers will also be required to frame policies to advertise the locations, health care facilities available and other information related to these PHCs so that the relevant information reach the target population, i.e. the low-income group women. With a combined effort of the government, policy makers, health care professionals, citizens and the women of reproductive age, the stigma of menstruation can be alleviated, and lives of menstruating women can be made easier and healthier.

## Data Availability

All data produced in the present work are contained in the manuscript

## Acknowledgements

The authors acknowledge Prof. Krutika B. Desai for her scientific inputs.

## Author Contributions

SS, SS, VS and MP conceived the study and designed the survey. SS, SS and VS conducted the survey and collected the data. SS, SS and VS tabulated the data and MP analysed the data. MP wrote the first manuscript draft. MP and PM reviewed and edited the manuscript. MP supervised the study.

